# Partner Dynamics at Maternal and New born Continuum of Care Enrollment among a Panel of Six Weeks Postpartum Women in Ethiopia, Community based Longitudinal Study; A Multinomial Logistics Regression Analysis

**DOI:** 10.1101/2024.06.05.24308524

**Authors:** Solomon Abrha Damtew, Mahari Yihdego Gidey, Fitsum Tariku Fantaye, Niguse Tadele Atianfu, Tariku Dejene, Kelemua Mengesha Sene, Tefamichael Awoke, Hailay G/kidan, Assefa Seme, Solomon Shiferaw

## Abstract

**Introduction:** In this era of SDG countries relatively achieving maternal and newborn health geographic coverage are moving to a new paradigm called completion of maternal and new born care continuum (MN- CoC). Measuring the influence of significant others: partner/or husband and community engagement are considered as pivotal and one of the pillar strategies to achieve of completion of MN-CoC. Unfortunately, there is a lack of policy framework for partner and/or husband involvement in pregnancy, childbirth and postnatal care and when exists there is a gap in policy and practice in developing countries context. Articulating and endorsing such policy is likely to promote husband and/or partner encouragement and support during at the enrollment to maternal and newborn care continuum (MN-CoC). Hence, quantifying the level of MN-CoC partner dynamics on antenatal care visit and/or discussion about where to deliver the index child at and identifying its correlates among a panel of six weeks postpartum women provides evidence for the policy articulation endeavor by the Federal Health Ministry of the Federal Democratic Republic of Ethiopian and developmental partners working on reproductive and maternal and newborn health care.

**Methods:** Community based nationally representative longitudinal data collected from a panel of pregnant and six weeks postpartum women were further analyzed. A total of 2,207 six weeks postpartum women who were married and/or living a partner were included in this analysis which was adequate to yield an unbiased estimates for MN-CoC partner dynamics. Multinomial logistics regression was run to identify correlates of partner Dynamics. Results were presented in the form percentages and odds ratio with 95% Confidence Intervals. Statistical significance was declared at p-value of 0.05.

**Results:** The proportion of partner and/or husband dynamics on MN-CoC of among six weeks postpartum women who reported that their partner and/or husband encouraged them to go to clinic for ANC and discussed with them about place of delivery for the index child was nearly 2/3 (64.67%; 62.21%, 67.04%).Besides, nearly one in 5 of them reported that their husband and/or partner did not encourage (18.2%; 16.64%, 20.33) and encouraged either of the first two MN-CoC two domains (17.08%, 15.25%, 19.10%). The region women residing, being in a polygamy marriage, contraceptive ever use history, attainment secondary/higher education and index child delivery place were found to contribute for the variation in MN-CoC partner dynamics.

The finding calls up on regionally sensitive activities and efforts with public-private partnership in service provision and targeting women with polygamy which in turn empower women to control over their fertility through increasing higher education enrollment, and diversifying access to contraceptive commodities are hoped to improve MN-CoC partner dynamics thereby enabling women in completing maternal and new born care continuum. Such endeavors and interventions are hoped to facilitate the ministry and other developmental partners’ comprehensive efforts to address the MN-CoC partner and/or partner dynamics in terms of policy articulation, advocacy, implementation, evaluation and revising it to fit its purpose and attain the desired targets.

**Author Plain English Summary:** *Purpose of Conducting the Study:* In every community, pregnancy and childbirth are expected to be joyful and positive experiences for the mother, the newborn and as well as for the families, however, neonatal, infant and maternal mortality is unacceptably high in low and middle income countries including Ethiopia. It is experienced not as the joyful event it should be, but as a dangerous and frightening time in their lives. In order to address such considerable problem, in the SDG era the focus of policy articulation, program concentration and research undertaking in maternal and newborn health care has shown a paradigm shift of measuring the completion of maternal, newborn care continuum (MN-CoC) by streamlining resources that were invested independently on each care continuum domains. Besides, WHO recommended completion of the three main domains of the maternal and newborn care continuum as pivotal strategy to improved maternal and new born health outcomes. Accordingly, partner encouragement, support and accompany during antenatal care, childbirth and postnatal care is one of the proposed strategy for pregnant women to be enrolled, retained and complete the maternal and new born care continuum including in sought care in the extended six postpartum period in particular and the inter pregnancy period in general. This is based on the evidence pool on the influence of significant others surrounding the women, notably; the community where they are residing and their partner and/or husband on health service use is considerably high. Determining the level of partner dynamics on the MN-CoC domains and identify its correlates is critical to track the progress of the proposed strategy. Nationally representative data collected from a panel of pregnant and six weeks the six post-partum women were used.

*Added Value of the Study:* Nearly 1 in 5 panel of women by their six week postpartum reported that they did not received any encouragement on the two first domains MN-CoC during their index pregnancy. Regional variation was observed in the level of partner dynamics at maternal and newborn care continuum enrollment domains (MN- CoC) and the variation was also explained by contraceptive ever use history as well. Women in polygamy marriage were less encouraged to go to clinic for ANC and lower opportunity to discuss where to deliver the index child with their partner and/or husband. .

*Implication of the Study:* The Federal Democratic Republic of Ethiopian Health Ministry and developmental partners need to articulate and endorse male involvement policy with region specific integrated public private strategies which improve women autonomy to control over their fertility and women higher education enrollment with a focus on women with polygamy so as to increase partner dynamics on MN-CoC. Partner encouragement on the first two domains of is key to enroll and retain pregnant women within the MN- CoC. There is a need to strengthen postpartum family planning counseling and diversifying the provision. Similarly women in polygamy needs attention. The need for installing preconception care in the health system to be provided in and around pregnancy and child birth; particularly the inter pregnancy preconception care package.

## Introduction

In the era of SDG were countries relatively achieving maternal and newborn health geographic coverage are moving to a new paradigm of completion of maternal and new born care continuum (MN-CoC) (1, 2). Completion of various maternal and new born care continuum (MN-CoC) is proved to improve maternal and new born health outcomes (3). Measuring the influence of significant others: community and partner/or husband is a considered as pivotal and one of the pillar strategies towards the level of completion of MN- CoC which is very critical to address the sustained higher level of maternal and neonatal mortality in lower and middle income countries in general and Sub Saharan Africa in particular (4).

Partner dynamics in maternal and new born care continuum is the involvement of husbands and/or partner in the pregnancy and child birth care (5, 6): consisting of encourage their wives to go to clinic for antenatal care (ANC), discuss and plan where to deliver and in seeking post natal care; companionship during antenatal care (ANC), accompany in his wife during child birth and postnatal care (7), taking part in the process of maternal and child health care continuum in particular and in reproductive health service use in general (4, 8). However, husband and /or partners misconceive this engagement as financial support and physical help alone (9) while overlooking their actual in person engagement, encouragement and discussion on maternal and health service use (10). WHO recommended that partner and/or husband involvement is a key strategy to improve uptake and completion of maternal and newborn care continuum (4).

Studies on male involvement showed that rate of male attendance at ANC with their pregnant wives was 69.4 in antenatal care showed that 69.4% and 57.00% male involved in antenatal care (6, 11) and delivery service use but only 23.1% entered ANC room with an overall involvement of 9% (12) while 66.2% involved in maternal, neonatal and child health care (13), however, these studies are based on cross sectional data and seldom addressed the emotional attachment of partner encouragement on ANC service use and discussion where to deliver during the index pregnancy which is very critical in accomplishing the maternal and new born care continuum (MN-CoC) while this study used a longitudinal data; merged baseline and six weeks postpartum, collected from cohort of pregnant and six week postpartum women. Such a robust evidence contributes much in filling the evidence gap to formulate policy framework for partner and/or husband engagement and encouragement during pregnancy, child birth and no and when exists there is a gap in policy and practice in developing country context (14–16).

In low and middle income countries such as Ethiopia where maternal, neonatal and infant mortality (17, 18) is unacceptably high, where most women prefer traditional birth attendant (19, 20), home delivery (21, 22) is very common, where the influence of the community on maternal and newborn health care continuum use has been shown considerable; measuring husband and/or partner influence is critical determinant in completing the of maternal and newborn care continuum (MN-CoC) (12, 16).

Evidences also shown that the active engagement of husband and/or partner ranging from encouragement and discussion along with serving as companion in matters related with pregnancy and child birth improves maternal and new born health outcome. According to WHO interventions to promote the involvement of husband and/or partner during pregnancy, childbirth and after birth are recommended to facilitate and support improved self-care of the woman, improved home care practices for the woman and newborn, and improved use of skilled care during pregnancy, childbirth and the postnatal period for women and newborns (5, 8).Despite such strong recommendation on husband and/or partner on involvement in his wife maternal and health service use, there is a lack of policy framework for husband and/or partner involvement in pregnancy, child birth and when exists there is a gap in policy and practice in developing countries context (1, 2, 4, 14, 23). Hence, articulating and endorsing such policy and strategy is pivotal to further improve maternal and newborn heath care continuum (MN-CoC) (16).

In the midst of this policy and strategy gap context, determining the magnitude of partner dynamics in terms of encouraging their wives during their pregnancy to go to clinic for ANC and discussing with them where to deliver the index pregnancy will provide evidence for the ministry of health and developmental partners to design and articulate strategies on husband and/or partner involvement along the maternal and newborn care continuum domains thereby improving maternal and new born health outcomes.

The objective of this study was to determine the level of partner and/or husband encouragement on the first two domains of the maternal and new born care continuum (MN-CoC) in Ethiopia and identifying the correlates contributing the variation in the encouragement.

## Methods and Data

### Sources Population and Study Design

Nationally representative merged cohort one baseline and the six weeks postpartum data collected from pregnant and six weeks postpartum women were used and in terms of follow up, it accounts half of the total follow up period, 12 or 13 months. The baseline and the six week follow up surveys were conducted form Nov 2019 to Jan 2021.The study was conducted in six regions of the country. The study employed a two stage stratified cluster sampling method. After the complete census was conducted in the selected enumeration areas all pregnant women with gestational age of 1 to 9 months who were either usual residents in the selected EAs or who came to the EA to deliver were enrolled and followed one year postpartum. Those who were within six weeks postpartum were also enrolled in the study. A total of **2,207** six weeks postpartum women were included in this analysis from a total of 217 enumeration areas. This analysis was restricted to women who were married or live together to better lean more on the influence of partner dynamics at enrolment of maternal and new born care continuum. The overall sample size and cell sample size adequacy was checked and found to be adequate enough in generating unbiased estimate for partner dynamics at maternal and newborn care continuum enrollment (MN-CoC).

### Field Work Procedure

Addis Ababa University’s School of Public Health in collaborative efforts with the Ethiopian Public Health Association, the Federal Health Ministry, Ethiopian Statistical Services, Bill & Melinda Gates Institute for Population and Reproductive Health (Johns Hopkins Bloomberg School of Public Health) were the implementers. The main sample units or enumeration areas (EAs) were chosen using the most recent frame to Ethiopia Population and Housing Census (PHC).

The first field work of the panel baseline survey was started by screening pregnant women and women who were less than 6 weeks post-partum. Data were collected at baseline and three follow up points, 6 week, 6 month and one year postpartum interviews. The details of field work implementation operations, sampling procedure, relevant methodological issues, measured indicators are were well articulated elsewhere. The protocol of PMA Ethiopia contains all the details on sample design and selection methods (24).

## Variables

### Outcome variable

The main outcome variable was partner dynamics at enrolment of maternal and new born care continuum (MN-CoC) among a panel of six week postpartum women. The following two question items asked women at the extended six week postpartum survey about their partner dynamics experience during the index pregnancy which is one of the criteria for their enrollment in this 2 years follow up study: 1. Partner encouraged you to go to clinic for antenatal care and 2. You & your partner discussed where to deliver (postpartum women). See the details of the question type and answer options and further recoding in the following table (**Table 1**).

**Table 1:** Items used to create the used to measure Partner Dynamic at maternal and new born continuum of care Enrollment among a Panel of Six Weeks Postpartum women, Baseline with Six Week Postpartum Data sets, Nov 2019 to Jan 2021.

|  | Variable | Question Items and Responses | Recode Outcome Category |
| --- | --- | --- | --- |
| Partner Dynamics at maternal | 1. Partner encouraged you to | 1. Yes<br>2. No, did Not discouraged<br>3. No, actively discourage<br>4. No partner | Four observation with no partner are excluded since this study restrict the study participants who are currently married and living as a partner. |
|  |  | -88. Do Not Know |  |
| and new<br>born<br>continuum<br>of care<br>Enrollment | go to clinic for<br>antenatal care<br>AND<br>2. You & your<br>partner<br>discussed<br>where to<br>deliver<br>(postpartum<br>women) | -77. partner not involved<br>0. No<br>1. Yes<br>**MN-CoC** Nominal composite variable was create with<br>the following categories:<br>0 partner do not encourage both MN-CoC domains,<br>base category<br>1 partner encourage either of the MN-CoC domains<br>2 partner encourage both MN-CoC domains | 15 observations (2 with DNK<br>response and 13 partner not<br>involved) were excluded from<br>the Analysis |

### Independent Variables

Independent variables were classified into individual-level variables and enumeration area-level variables. Individual-level independent variables further categorized into socio-demographic/economic characteristics variables, parity and other RH characteristics and contraception ever use were considered in the study.

Group or enumeration area (EA) level variables included two integral variables namely, region and place of residence while two derived EA level variables, EA level wealth derived from the household level wealth, while proportion of women who completed secondary or above educational level per EA were derived from the respective women and husband/partner educational status. “Region” was grouped into six categories, 1=Tigray 2 = Afar, 3= Amhara, 4=Oromia, 7= SNNPRs and 10= Addis Ababa city administration. Place of residence follows the default urban/rural classification.

Health service use during the index pregnancy and in the post-partum period such as antenatal care seeking from health professional, HEW and place the index child delivered, birth attendant of the index child, starting using contraceptive by six week postpartum and contraceptive ever us history are also include as covariates in this this study.

### Analysis and Measurement

A merged cohort one data consisting baseline data and six week data from cohort one PMA survey conducted from Nov 2019 to Jan 2021 were used for this study (24). Stata v16 was used for this analysis. Frequencies and percentages were computed to characterize the study population. Chi-square test statistics was computed to check cell sample size adequacy and the sample size was found to be adequate to provide unbiased estimates.

Exploratory data analysis was run for data cleaning thereby checking item nonresponse rate for every variable considered in this analysis and don’t know response which were later excluded from the analysis. Following this variable were recoded to create biological plausible categories along with checking distribution of the recoded variables using mean and proportion as appropriate. No sign of multicollinearity detected among variables in the final model. Multinomial logistics regression was used to identify important predictors of partner dynamics at maternal and new born care continuum (MN-CoC). At bivariate analysis a p value cut off point of 0.25 was used to select candidate variable for multinomial multivariable logistics regression analysis. Results were presented in the form of percentage, and odds ratio with 95% CI. Significance was declared at a significance level of 0.05. Results were reported based on weighted count.

### Data Quality Management and Control

PMA Ethiopia data were collected using standard and pretested tool which was translated in to three local languages (Tigrigna, Afan Oromo and Amharic) after the provision of hands-on intensive ToT, RE training with mock interviews. Close supervision during filed work, timely progress report and hierarchal errors correction, 10% random check with dedicated re interview form were some of the modalities used to maintain the quality of the collected data, the detail is reported somewhere else (24). Data completeness for variables and items for creating composite variables was checked by exploratory analysis following which any item nonresponse was excluded from the analysis. Frequency run to exclude responses with do not know (DNK) and no response (NR).

### Ethical consideration

This study involved a secondary analysis of de_identified data from the PMA Ethiopia. The PMA Ethiopia survey was conducted strictly under the ethical rules and regulations of world health organization and IIRB of Ethiopian Health and Nutrition Research Institute (EHNRI). Informed consent was obtained from respondents during the data collection process of PMA Ethiopia on data collection on Nov 2019 to Jan 2021. PMA surrey has been also conducted after obtained ethical approval from the College of Health Sciences at Addis Ababa University and Bloomberg School of Public Health at Johns Hopkins University in Baltimore, USA. PMA_ETH Publicly available Cohort one baseline and six weeks postpartum datasets were accessed after submitting a concept note for this piece of specific work form the PMA data cloud server archive via. https://www.pmadata.org/data/request-access-datasets.

Since this study involves analysis of already collected secondary data there is no need to consent, rather, concept note was submitted to get permission for data use.

“Minors less than 15 years as per the law were not included in this study. Informed verbal consent was take from study participants.” Moreover, women of reproductive age group or women child bearing age were include in the study. The survey includes topics on related on family planning, sexual history and other reproductive health issues which is declared are right of women by international declarations and as supported by evidence: Rimon JGII, Tsui AO. Regaining momentum in family planning. Global Health Science Practice. 2018; 6(4):626–628. https://doi.org/10.9745/GHSP-D-18-00483. 2018 and related documents. Hence, asking consent another person or guardian for women girls 15 to 17 years will contradict with waived room to direct ask the herself about her sexual and reproductive issues. Moreover standard surveys including Demographic and health surveys include women of child bearing age as this study did.

## Results

### Magnitude of Partner Dynamics at Maternal and Newborn Continuum Care Enrollment (MN-CoC) among a panel of six weeks postpartum Women

This study generated findings on partner dynamics on the first two domains of maternal and newborn care continuum (MN-CoC) among a panel of women at their six weeks postpartum. This study reported the proportion of partner dynamics at the maternal and newborn care continuum enrollment (MN-CoC) as measured by creating a composite variable called partner dynamics from the two question items asked namely: women reported that her husband and/or partner encouraged her to go to clinic for antenatal care (ANC) and discussed with them where to deliver the index child. Three categories were created from these two domains: do not encourage both the first two domains, encouraged either of the first two domains and encourage both the first two domains.

The proportion of partner and/or husband dynamics on MN-CoC of among six weeks postpartum women who reported that their partner and/or husband encouraged them to go to clinic for ANC and discussed with them about place of delivery for the index child was nearly 2/3 (64.67%; 62.21%, 67.04%).Besides, nearly one in 5 of them reported that their husband and/or partner did not encourage (18.2%; 16.64%, 20.33) and encouraged either of the first two MN-CoC two domains (17.08%, 15.25%, 19.10%). **(Fig 1).**

**Fig 1:**
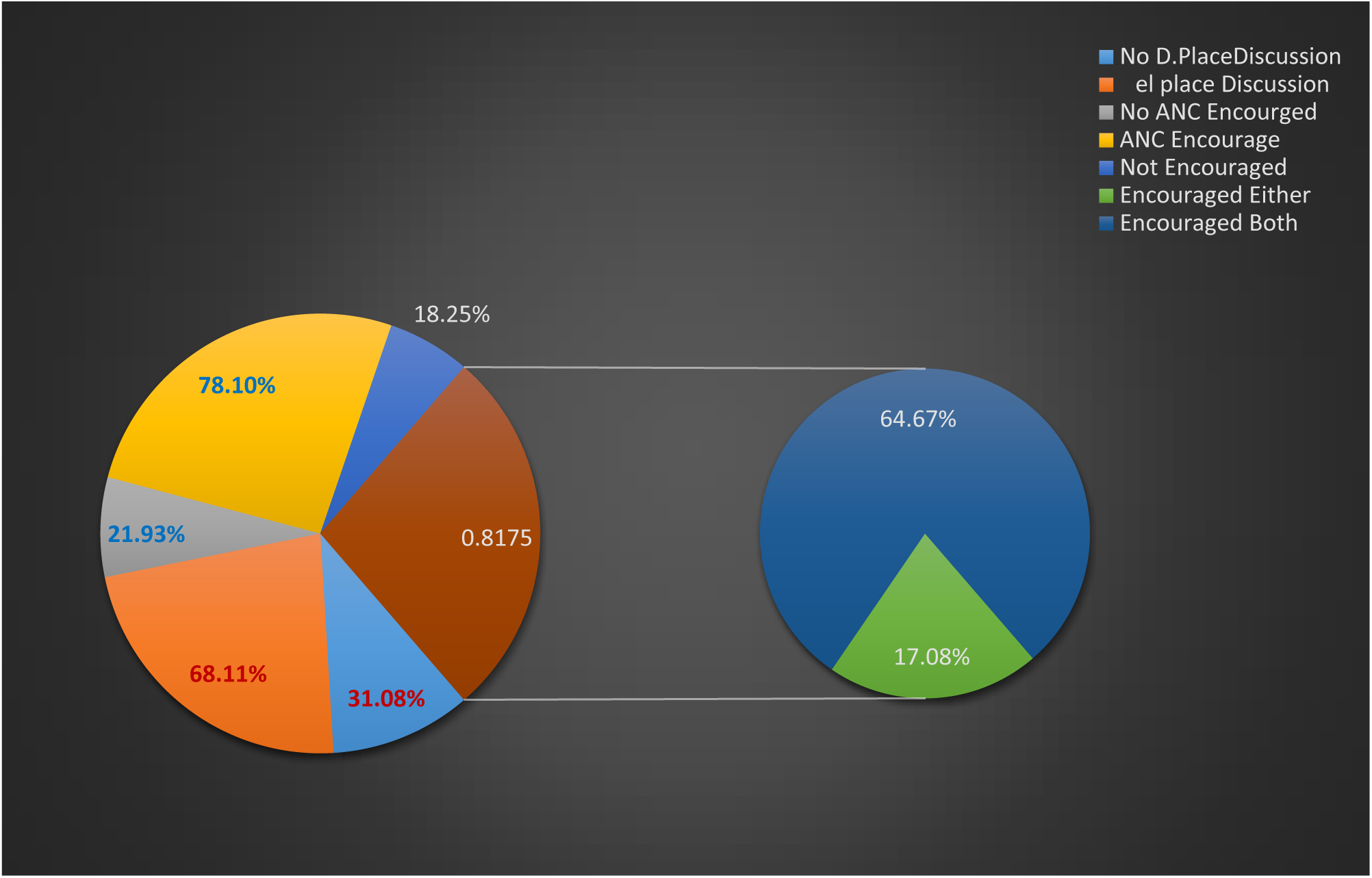
Partner Dynamics at maternal and new born continuum of care Enrollmentamong a Cohort of Women and its correlates in Ethiopia

The proportion of a panel of six weeks postpartum who reported that their husbands and/or partner encouraged them to go to clinic for ANC was 78.10% (75.90%, 80.01%) while 68.11% (65.70%, 70.44%) six week post-partum panel women reported that they have discussed with their partner and/or husband where to deliver the index child during the index pregnancy (**Fig 1).**

### Distribution of Partner Dynamics at Maternal and Newborn Care Continuum Enrollment among a Cohort of Women, Evidence from PMA Cohort One and Six Weeks Postpartum Surveys

This study generated findings on partner Dynamics for a panel of women at their six weeks postpartum follow up. Partner dynamics among a panel of six weeks postpartum women at the maternal and newborn continuum care enrollment (MN-CoC) was measured as the composite of the first two domains of the maternal and newborn care continuum: women reported that her husband and/or partner encouraged her to go clinic for ANC and discussed with her where to deliver about the index child during their index pregnancy **(Table 1).**

This study identified correlates of partner dynamics at maternal and new care continuum enrollment (MN- CoC) for the partner encourage both of the first two domains, encourage either of first two domains and did not encouraged both first two domains. In absolute terms the partner dynamics among a panel of six weeks postpartum women showed variation across the selected covariates **(Table 2).**

**Table 2:**
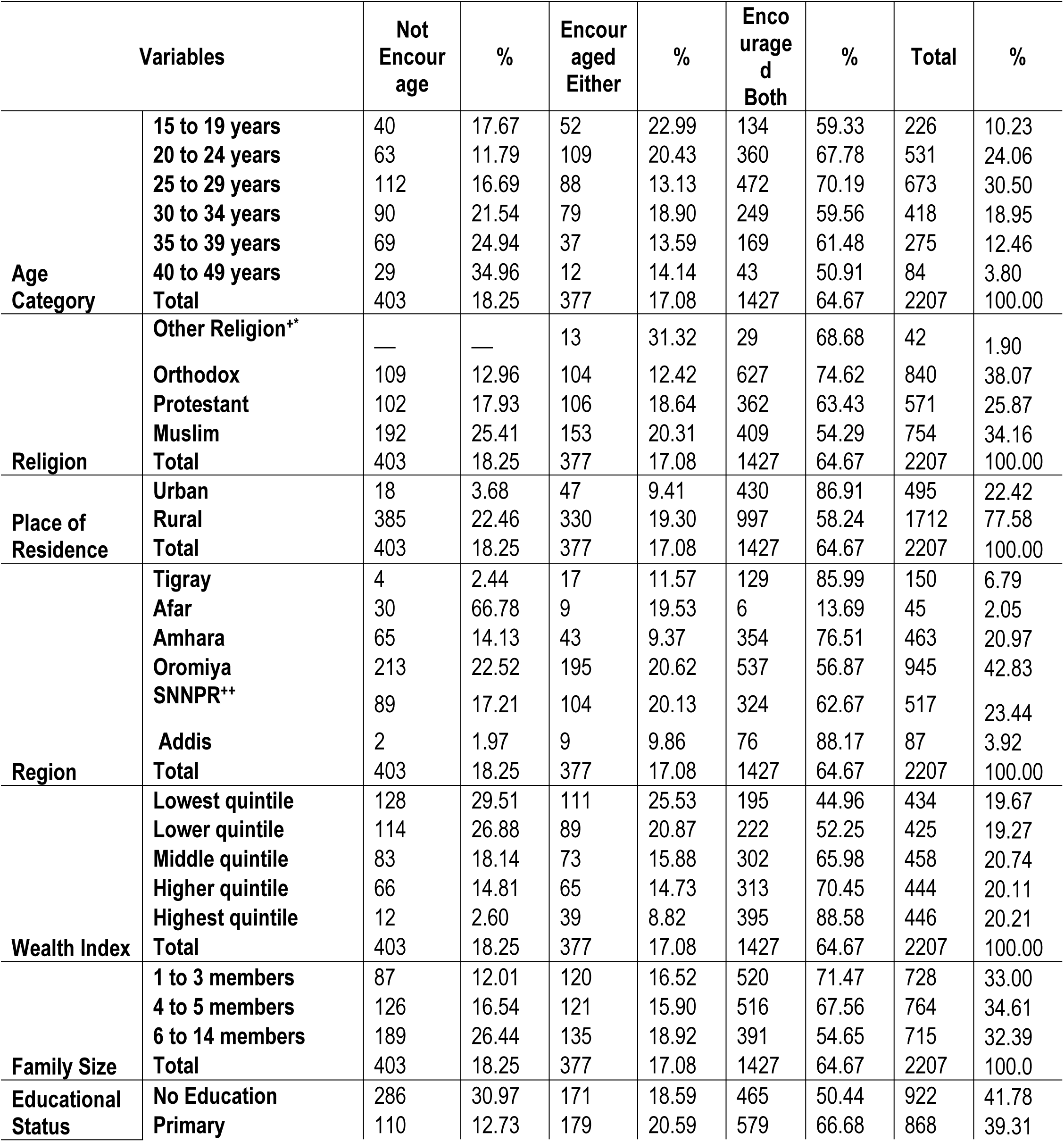
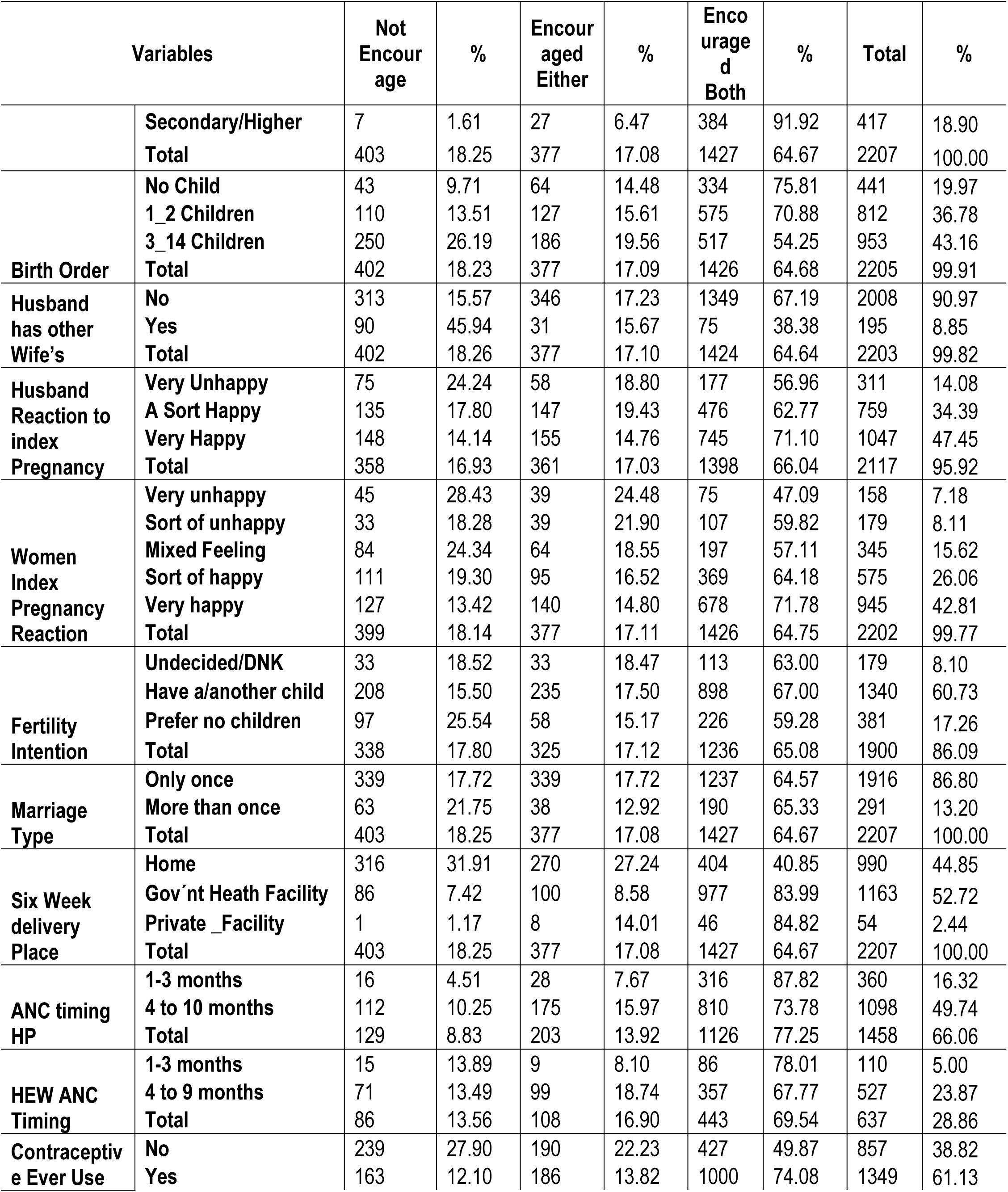
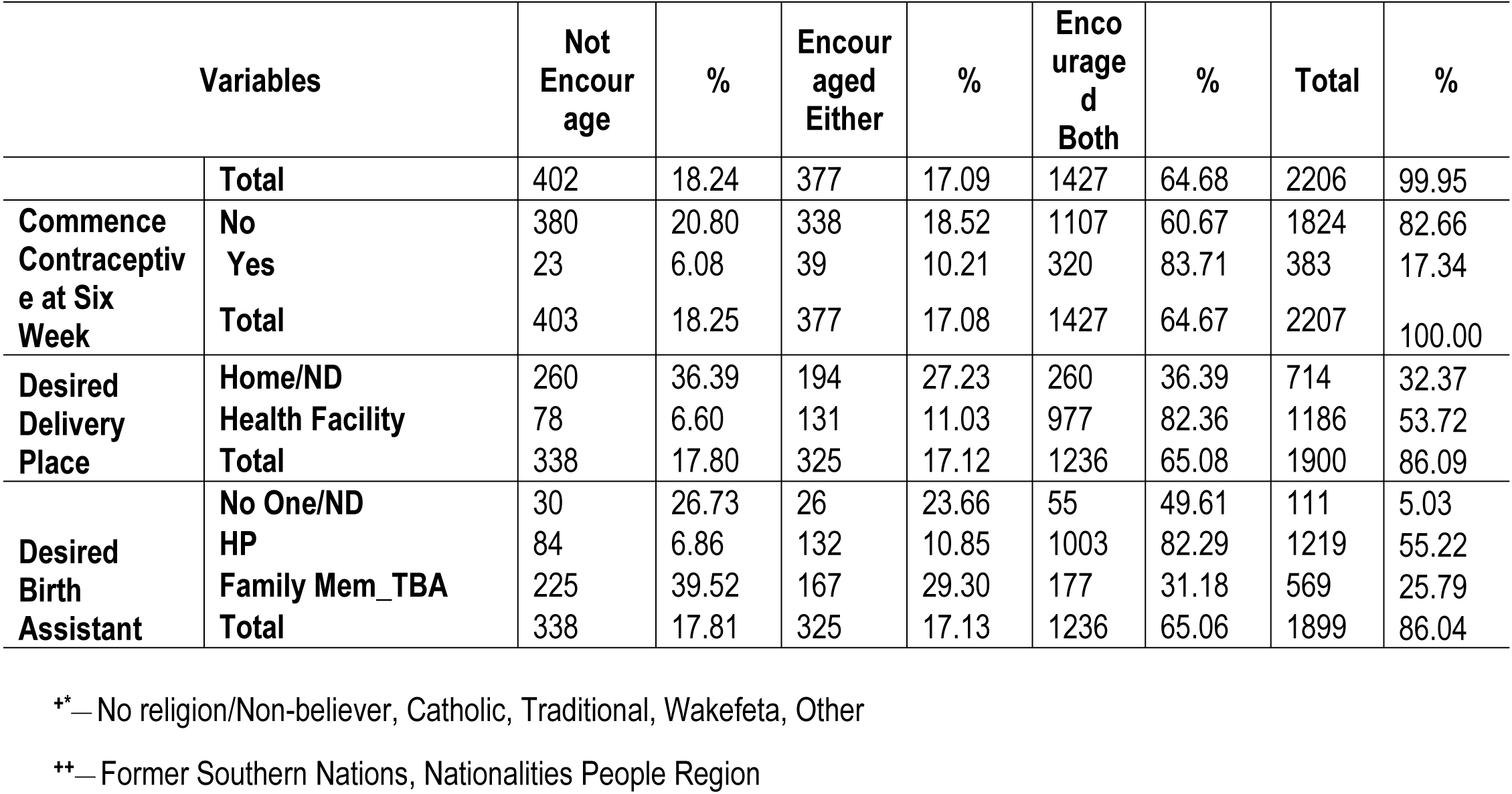
Partner Dynamics at maternal and new born continuum of care Enrollment among a Cohort of Women in Ethiopia and its correlates in Ethiopia, Community based Follow Up Study; evidence from PMA Cohort One Merged Baseline and Six Week Postpartum Data Sets, Nov 2019 to Jan 2021 (n= 2,207)

The partner dynamics (MN-CoC) showed an increasing pattern with age for the did not encouraged category of the outcome, accordingly, among women in the elders (40 to 49 age category), one third (34.96%), 14.14% and half (50.91%) reported that their husband and/or partner do not encourage both domains of the partner dynamics; encourage either to go to clinic for ANC or discuss about place of delivery for the index child; and partner encourage both domains of partner dynamics at the maternal and newborn care continuum enrollment. For Muslims religion followers, this respective figure was 25.41%, 20.31% and 54.29% while no women reported that their partner did not encouraged for the other religion followers. Similarly, among urban residents 3.68%, 9.41% and 86.91% reported that their partner and/or husband did not encourage, encourage either and encourage both of the first two MN-CoC domains respectively. Likewise, this same figure stood 2.44%, 11.57% and 85.99% for residents of Tigraye and 14.13%, 9.37% and 76.51% for residents of Amhara regions **(Table 2).**

In terms of wealth quintile, the proportion of women from the well to do households who reported that their partner and/or husband did not encouraged, encourage either of the two domains and encouraged both the first two MN-CoC domains was 2.60%,8.82% and 88.58% respectively. This same figure was 26.44%, 18.92% and 54.65% for those living with a larger family size of 6 or more. Similarly, the proportion for the respective MN-CoC partner dynamics domains among those women with no formal education stood for 30.97%, 18.59%, 50.44% respectively **(Table 2).**

The proportion of six weeks postpartum women who reported that their partner and/or husband do not encouraged, encouraged either domains and encouraged both domains of the partner dynamics among those with higher birth order was 26.19%, 19.56% and 54.25% respectively. This proportion was 18.52%, 18.47% and 63.00% for those who have not decided to have an additional child. In a similar fashion, for those reported who felt happy when learned their index pregnancy, this figure stood for 13.42%, 14.80% and 71.78% respectively while the respective proportion was 24.24%, 18.80% and 56.96% of for women reported that whose husbands felt unhappy when learned about the index pregnancy. Similarly, the respective partner dynamics proportion for women who were married more than once was 21.75%,12.92% and 65.33% respectively while the respective figure for those whose husband had other was 18.26% 17.10% and 64.64% respectively **(Table 2).**

For those who delivered the index pregnancy at the government health facilities, 7.42%, 8.58 and 83.99% of them reported that did not received encouragement, receive either domain encouragement and both domain encouragement from their partner on the first two MN_ CoC partner dynamics domains while for those who received their first ANC from professional heath care provider other than health extension workers (HEW) during the first trimester the respective proportion was 4.51%, 7.67% and 87.82% for the respective partner dynamics domains. The respective figure for those who obtained their first visit in the first trimester form HEW, was 13.89%, 8.10% 78.01%. Besides among those who started to use contraceptive at six weeks post-partum, 83.71% reported that their partner encouraged to go to clinic for ANC and discuss where to deliver; 10.21% reported either of the two partner dynamics while 6.08% reported that got encourage neither of the first two MN-CoC domains.

This MN-CoC partner dynamics figure for those who have ever used contraceptive commodities was 74.08%, 13.82% and 12.10% respectively. Besides, among those whose desired place of delivery for the index was home/not decided 1/3 (36.39%) reported that their partner never encourage and encouraged both domains while 27.23% of them reported that their husband encourage either of the partner dynamics domains. Similarly, for those whose desired birth assistant for the index child was no one or did not decided 26.73%, 23.66% 49.61% reported that their partner did not encourage, encourage either of the two and encourage both of the partner dynamics domains at maternal and newborn care continuum enrollment respectively **(Table 2).**

### Regional variation of Partner Dynamics at Maternal and Newborn Care Continuum, for both domains encouraged category, evidence from PMA cohort 1 Baseline and Six weeks postpartum merged Data

As can be seen in figure 2 below, the level of partner dynamic as measured in terms of the encouragement for both MN-CoC domains: to go to clinic for antenatal care and discuss where to deliver the index child showed substantial regional variation from 13.69% among residents of Afar region to 88.17% in Addis Ababa. The MN-CoC regional variation raged from 1 in 7 women among residents of Afar region to 1 in 2, higher than 7 in 10, close to 9 in 10 and 6 in 10 among residents of Oromiya, Amahara, Tigiraye and the former SNNPR regions respectively **(Fig 2).**

**Fig 2.**
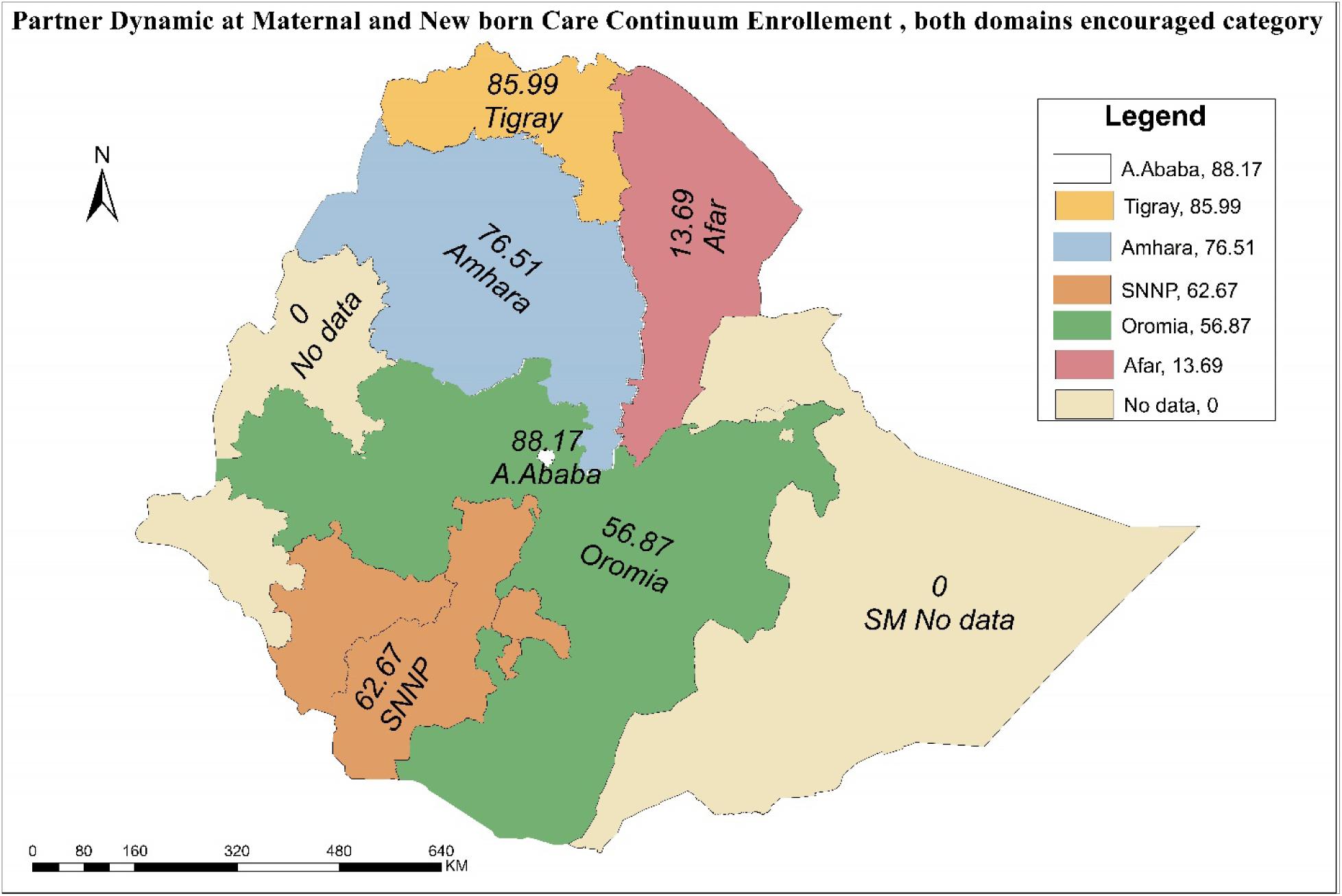
Partner Dynamics at maternal and new born care continuum Enrollment, for both domains encouraged category, evidence PMA cohort 1 Baseline and Six weeks postpartum Data. (Source: updated shape file obtained from humanitarian data exchange https://data.humdata.org)

### Correlates of Partner Dynamics at maternal and new born care continuum Enrollment among a Cohort of Six Weeks Postpartum Women in Ethiopia, Community based Follow Up Study

This study identified correlates of partner dynamics at maternal and new care continuum of enrollment (MN- CoC), in terms of encouragement their pregnant wives to go to clinic for ANC and discuss where to deliver during the index pregnancy. This study identified and reported correlated of partner dynamics for the composite variable created: encourage either domains and encourage both domains using do not encourage both as a base outcome category in the multinomial regression modeling procedure

The finding showed that there is regional variation in partner dynamics towards maternal and new born care continuum enrollment (MN-CoC) and variation by whether husband has other wives and women educational attainment as well. It also showed variation by the place the women gave birth of the index child and by contraceptive use history (explain variation for the either encourage category alone) (**Table 3).**

**Table 3:**
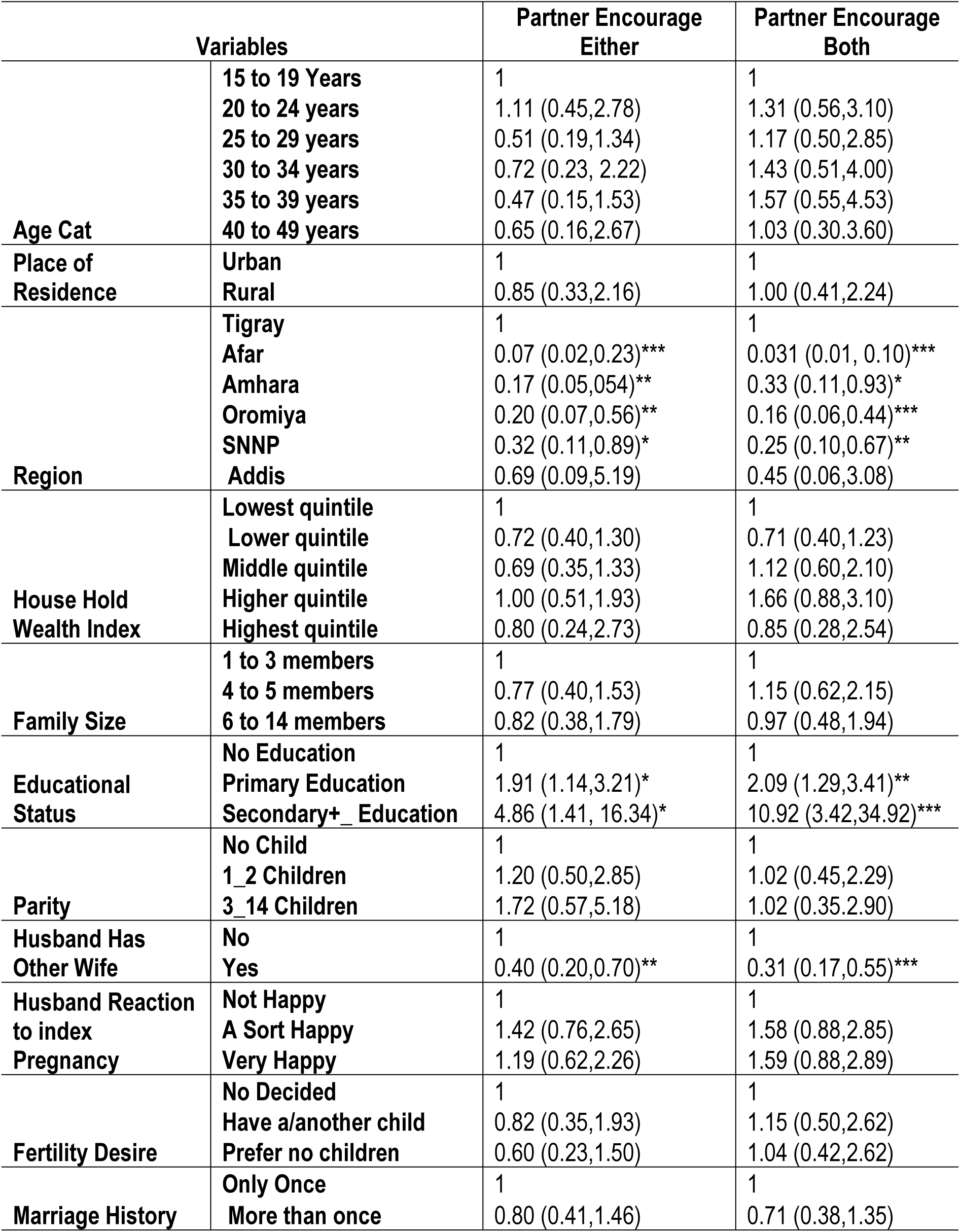
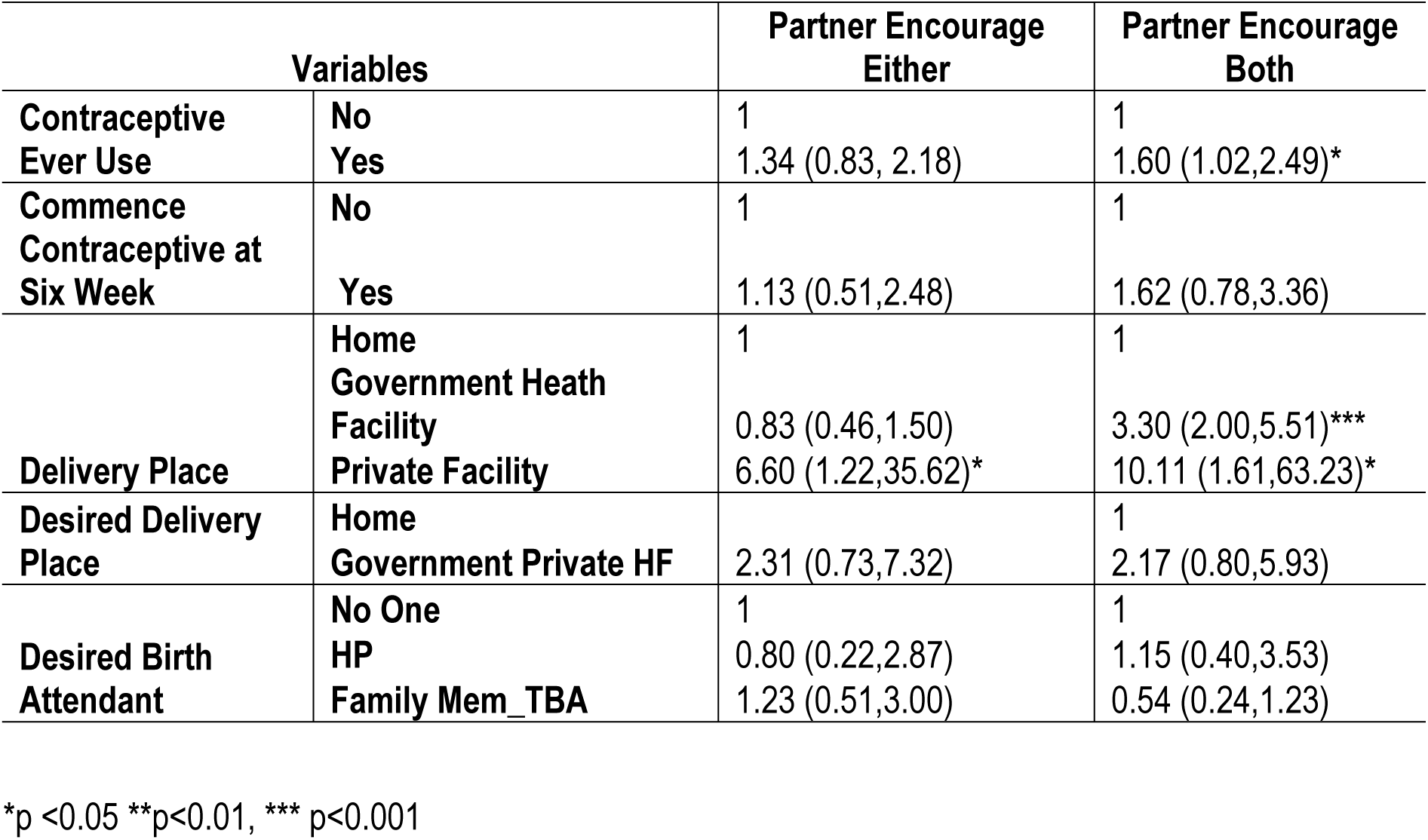
Correlates of Partner Dynamics at maternal and new born continuum of care Enrollment among a Cohort of Women in Ethiopia and its correlates in Ethiopia, Community based Follow Up Study; A multinomial Logistics regression Analysis, Nov 2019 to Jan 2021.

The finding showed that there is regional variation in partner dynamics (MN-CoC). The likelihood of partner dynamics (MN-CoC encouraged both domains) at maternal and newborn health care continuum enrollment among women who are from Afar, Oromia and the former SNNPR Regions was (ARRR: 0.07 (0.02, 0.23)), (ARRR: 0.20 (0.07, 0.56)) and (ARRR: 0.25 (0.10, 0.67)) times lower respectively. Similarly, those women living with a husband and/or partner who had other wives had 69% ((ARRR: 0.31 (0.17, 0.55)) lower odds of partner encouragement of both of the partner dynamics domains at maternal new born care enrollment (**Table 3).**

On the other hand, the likelihood was (ARRR: 3.30 (2.00, 5.51)) and (AOR: 10.11 (1.61, 63.23)) times higher among six weeks postpartum women who reported that they have gave birth of their index child in governmental and private facilities respectively. Similarly, this likelihood was (ARRR: 1.60 (1.02, 2.49) times higher among women who were contraceptive ever users compared with life time non users. Besides, women with primary education and secondary plus educational attainment had (ARRR: 2.09 (1.29, 3.41)) and (10.92 (3.42, 34.92)) times higher likelihood of the MN-CoC partner encouragement of both of the partner dynamics domains at maternal new born care enrollment compared with their non-educated counterparts **(Table 3).**

For partner encouraged either of the domains, the finding showed that there is regional variation in partner dynamics on MN-CoC. The likelihood of partner dynamics (encouraged either domains) at maternal and newborn health care continuum enrollment among a panel of six weeks postpartum women was (ARRR: 0.031 (0.01, 0.10)), ((ARRR: 0.16 (0.06, 0.44)) and Amhara ((ARRR: 0.17 (0.05, 054)) times lower in Afar, Oromia and Amhara regions respectively. Similarly, those women living with a husband and/or partner who had other wives had 60% ((ARRR: 0.40 (0.20, 0.70)) lower odds of partner encouragement of either of the partner dynamics domains at maternal new born care enrollment **(Table 3).**

On the other hand, women who achieved primary education had (ARRR: 1.91 (1.14, 3.21)) and secondary plus education had ARRR: (4.86 (1.41, 16.34)) higher odds. Besides those who gave birth of their index child at private facilities (AOR: 6.60 (1.22, 35.62)) times higher likelihood of partner encouragement on either of the partner dynamics domains at maternal new born care enrollment **(Table 3).**

## Discussion

Maternal and new born health outcomes can be greatly improved with cost effective intervention among others completing the maternal and newborn care continuum plays pivotal role in improving maternal and newborn health outcomes. Besides, in midst of the SDG era there is a paradigm shift in terms of policy articulation, service provision and research focus on maternal and new born health service; which calls service provision as continuum of maternal and new born health care which call all pregnant women enrolled to ANC care should be retained to get there recommended cares namely; ANC4+ and skilled birth care and immediate post natal care. This is not achievable without the involvement of partner and/or husband. Evidences showed that one of the bottlenecks for such the persistent poor maternal and fetal outcome despite the visible efforts in maternal and child health (MCH) service provision is scarce partner and/or husband involvement and encouragement on their wives to be enrolled and retained in the maternal an newborn continuum of care to get the recommended three domains of the care continuum.

In the midpoint of the SDG era and at this time when modern primary health care services are being available and accessible near to the community to a larger extents; and the government of Ethiopia has been showing very committed to provide delivery care, most antenatal and postnatal care services with exemption from charge; and most importantly being at the era when the role of husbands involvement and the community support on maternal and newborn health service use has been catching policy makers, program managers and researchers attention and their direction of focus; documenting the proportion of partners and/or husbands who encouraged their wives to go to clinic for ANC use and discussing with their wives where to deliver the index child and identifying the correlates contributing for the variation of such partner dynamics is very critical in availing actionable evidence to improve maternal and new born outcome by increasing skilled delivery care at health facilities. Therefore, this study focused on determining the level of partner dynamics which was measured as ANC use encouragement and discussing where to deliver the index child and identified its correlates by further analyzing a follow up data from PMA for action Ethiopia.

In addition, to the conventional discussion method, the authors mainly write the discussion using an indirect method of discussion such us comparing the finding with accomplishment goals stated in the national and international relevant policy and program endorsed documents and targets such as the success and challenge of the Ethiopian health extension program, the national Health sector transformation plans.

The finding that nearly two third (64.67%; 62.21%, 67.04%) of six weeks postpartum women reported that their husband and/or partner encouraged them to go to clinic for ANC and discussed with them about place of delivery for the index child, while nearly one in 5 women reported that their husband and/or partner not encourage the first two MN-CoC partner dynamics domains (18.2%; 16.64%, 20.33) and either encouraged either of the two (17.08%, 15.25%, 19.10%). This considerable proportion in partner dynamics in terms of encouragement to go to clinic for ANC and discuss where to deliver the index child might be related with the success of health extension program (HEP) in Ethiopia, Health Sector Transformational Plans I_II (HSTP 2) (18, 25, 26) and the endorsement and implementation of the new reproductive health strategy running 2021 to 25 in which preconception care related targets were embedded (25). The finding nearly one third of the six week postpartum did not received encouragement on both MN-CoC partner dynamic domains might be related women experiencing low quality antenatal care (ANC) and did not seek the recommended four or more ANC visits during their most recent pregnancy and childbirth (26) in which case less likely to received counseling on the role and involvement of their husband their care during pregnancy and childbirth ; a recent study based entitled “Effective coverage of antenatal care services in Ethiopia: a population-based cross-sectional study. BMC Pregnancy Childbirth. 2024;24(1):330” indicated that the overall effective ANC coverage was low, primarily due to a considerable drop in the proportion of women who completed four or more ANC visits.

To this end, the implication of the finding that nearly two third (64.67%; 62.21%, 67.04%) of women reported that their husband and/or partner encouraged them to go to clinic for ANC and discussed with them about place of delivery indicated that Ethiopia has a long way to go to escalate partner encouragement and engagement along the maternal and new born care continuum care (4, 8). Furthermore, the expansion of the urban Health extension professional and addition of level IV Health extension workers in rural set up should be used as a stepping stone to improve the provision and completion of care for maternal and newborn care continuum (27) at community level thereby enabling women to get informed counseling about their role of husband and/or partner encouragement and involvement at maternal and new born care continuum enrollment thereby improving continuum of maternal and new born care. Moreover, the government’s effort to improve quality of maternal and child health care and respectful maternity care (28) along with the continuum of care is likely to encourage women to plan their pregnancies and subsequently to be to be encouraged by their husband and/or partner and the variation in community support for pregnant and postpartum women to utilize the three domains of the maternal and new born continuum of care packages also contributes for this observed variation in the partner dynamics: Community based 2 year Cohort follow up Study (29) as well as lack of partner encouragement that his wife to clinic for ANC, discuss where to deliver and seek postnatal care for the index pregnancy as reported by a study (30) entitled “”The impact of partner autonomy constraints on women’s health-seeking across the maternal and newborn continuum of care.”” : the fact that nearly one in 5 (18.2%; 16.64%, 20.33%) women reported that their husband and/or partner not encourage to go to clinic for ANC and is in line with a study (30).

The regional variation in the finding that partner dynamics among six weeks postpartum women might be related with the varying degree of birth preparedness and complication readiness activities across regions and better implementation of the health sector transformation plans in general and the reproductive health policy running 2021 to 25 in particular (18, 25, 31). The observed regional variation might be further related with the engagement and support of developmental partners working with the Federal Health Ministry Health and Regional health bureaus in terms of direct support in maternal and reproductive commodities provision by push strategy and research fund support. The variation in provision of male friendly service (32) across regions along with sociocultural and religious influence on male involvement and/or encouragement might also contribute for the observed regional variation (33).

The finding that educational attainment increased the likelihood of partner dynamics was in line with the assertion that attending higher education tend to improve women reproductive autonomy (34). Education increase partner dynamics in terms of encouraging women to go to clinic for ANC and discuss place of deliver for the index child might be related with educated women are like to have persuasive ability and have good communication skill, hence, resulting a good couples communication (35) and showing the role of women decision making ability on their perception of their status in the family (36).

In line with a study (37) type of health facility where the index child delivered was found to be related with improved partner dynamics: nearby health facility increase male involvement and in this study place of delivery being health facility increased the likelihood of partner dynamics, it might also be related with proximity to health facilities to get delivery service and receive reproductive commodities (38). It might also be re related with getting quality delivery care since the government of Ethiopia has given due attention on quality of care provide in recent years (39). It might also be related that such women are aware of their reproductive service use right (40, 41) making the partner and/or husband to engage his wife in matters related with reproductive health including encouraging her to go to clinic for ANC and discuss where to deliver and as part of couples birth preparedness and complication readiness (42).

Contraceptive ever use only increase the likelihood of partners dynamics might be emanated from women alone decision making power on contraceptive commodities use (43–48) and reproductive service use autonomy (49) which enable women to aware of their reproductive rights and health service need alarming their partner and/or husband to encourage them to go to clinic for ANC and discuss with them about where to deliver the index child as part of couples birth preparedness and complication readiness (42).

The finding that husbands has other wives lower MN-CoC partner dynamic on the first two domains was mentioned in a study entitled “The practice of polygamy on the utilization of reproductive health services among married women in Ghana” reported that women have lower likelihood of maternal health service namely; the recommended antenatal care visit and skilled birth uptakes. Besides the study recommended interventions on reproductive health may need to priorities women in polygamous marriages in order to improve the utilization of skilled ANC, assisted skilled birth, and modern contraceptive services (50). Another study showed that polygamous marriage had psychological sequel on women and children health outcomes (51).

## Conclusions

The proportion of partner and/or husband dynamics on MN-CoC of among six weeks postpartum women who reported that their partner and/or husband encouraged them to go to clinic for ANC and discussed with them about place of delivery for the index child was nearly 2/3. Moreover, nearly 1 in 5 women reported that their husband and/or partner did not encourage (18.2%; 16.64%, 20.33%) and either encouraged either of the two MN-CoC domains (17.08%, 15.25%,19.10%) calls up on key actors to design and implement regionally sensitive collaborative public private partnership activities and efforts that empower women through secondary or higher education enrollment and which also addressed polygamy are hoped to improve husband and/or partner involvement in their wives pregnancy and child birth in general and maternal and new both care continuum service use encouragement in particular. Contraceptive use history is also another source of variation for partner dynamics of the both encourage category, hence, diversifying access to contraceptive commodities is likely to help the ministry and other developmental parents to address the husband and/or partner MN-CoC dynamics in terms of policy articulation, advocacy implementation, , evaluation and revising it to fit its purpose and attain the desired targets.

## Data Availability

The datasets generated during the study are publicly available from the PMA website. https://www.pmadata.org/data/request-access-datasets.

## List of Abbreviations

ARRR: Adjusted Relative Risk
MN-CoC: Maternal and Newborn Care Continuum
EA: Enumeration Areas
HEW: Health Extension Worker
HH: households
PMA: Performance for Monitoring for Action Ethiopia
RE: Resident Enumerator
SNNPR: Southern Nations, nationalities and Peoples Region.
ToT: Trainer of Trainees.

## Declarations

### Ethics approval and consent to participate

This study involved a secondary analysis of deidentified data from the PMA Ethiopia. The PMA Ethiopia survey was conducted strictly under the ethical rules and regulations of world health organization and Research Ethics Review board of the College Health Sciences and Medicine of Addis Ababa University. Informed consent was obtained from respondents during the data collection process of PMA Ethiopia on the baseline data collection on Oct 2021. Minors less than 15 years as per the law were not included in this study. Informed verbal consent was take from study participants. PMA surrey has been also conducted after obtained ethical approval from Bloomberg School of Public Health at Johns Hopkins University in Baltimore, USA.

### Consent for publication

N/A not applicable

### Competing interests

The authors declare that they have no competing interest.

### Funding

The authors did not obtained any funding.

## Author Contributions

**Conceptualization**: Solomon Abrha Damtew.

**Data curation**: Solomon Abrha Damtew, Tesfamichael Awoke, Tariku Dejene.

**Formal analysis**: Solomon Abrha Damtew.

**Investigation**: Assefa Semen, Solomon Shiferaw.

**Methodology**: Solomon Abrha Damtew,

**Software**: Solomon Abrha Damtew, Tariku Dejene

**Supervision**: Assefa Semen, Solomon Shiferaw.

**Funding acquisition and Resources:** Assefa Seme, Solomon Shiferaw.

**Project administration**: Solomon Abrha Damtew, Mahari Yihdego Gidey,Niguse Tadele Atinafu, Hailay G/Kidan

**Validation**: Solomon Shiferaw, Assefa Seme.

**Visualization**: Solomon Abrha Damtew, Fitsum Tariku Fantaye, Tariku Dejene.

**Writing – original draft:** Solomon Abrha Damtew.

**Writing – review & editing**: Solomon Abrha Damtew, Tariku Dejene, Fitsum Tariku Fantaye, Tesfamichael Awoke, Mahari Yihdego Gidey,Niguse Tadele Atinafu, Hailay G/Kidan Kelemua Mengesha Sene, Assefa Seme, Solomon Shiferaw.

All authors reviewed and approved the final version of the manuscript.

## Acknowledgement

The authors are very thankful for Performance Monitoring for Action (PMA) Project. The very cooperative respondents and dedicated field team deserves a heartfelt acknowledgement. We would like to extend our heartfelt appreciation to staff of all Ethiopian Red Cross Training Center Kereyu resort_Adama for their hospitality in facilitating PMA staff trainings Staff and for hosting two round PMA large scale data analysis and use trainings. Last but not least the trainers from AAU and JHU deserve recognition as well as Linnea A.Zimmermańs mentorship following the training and fellow trainees’ inspiration.

## Notes

### Competing Interest Statement

The authors have declared no competing interest.

### Clinical Trial

NA

### Funding Statement

The authors did not receive any funding for this piece of work.

### Author Declarations

Ethical consideration Since this study involves analysis of already collected secondary data there is no need to consent, rather, concept note was submitted to get permission for data use. “Minors less than 15 years as per the law were not included in this study. Informed verbal consent was take from study participants.” Moreover, women of reproductive age group or women child bearing age were include in the study. The survey includes topics on related on family planning, sexual history and other reproductive health issues which is declared are right of women by international declarations and as supported by evidence: Rimon JGII, Tsui AO. Regaining momentum in family planning. Global Health Science Practice. 2018 6(4):626–628. https://doi.org/10.9745/GHSP-D-18-00483. 2018 and related documents. Hence, asking consent another person or guardian for women girls 15 to 17 years will contradict with waived room to direct ask the herself about her sexual and reproductive issues. Moreover standard surveys including Demographic and health surveys include women of child bearing age as this study did. To this end the primary data was conducted under standard ethical clearance consideration after getting Ethical clearance approval from the College of Health Sciences at Addis Ababa University and Bloomberg School of Public Health at Johns Hopkins University in Baltimore, USA. This study involved a secondary analysis of de_identified data from the PMA Ethiopia. The PMA Ethiopia survey was conducted strictly under the ethical rules and regulations of world health organization and IIRB of Ethiopian Health and Nutrition Research Institute (EHNRI). Informed consent was obtained from respondents during the data collection process of PMA Ethiopia on data collection on Nov 2019 to Jan 2021. PMA surrey has been also conducted after obtained ethical approval from the College of Health Sciences at Addis Ababa University and Bloomberg School of Public Health at Johns Hopkins University in Baltimore, USA. PMA_ETH Publicly available Cohort one baseline and six weeks postpartum datasets were accessed after submitting a concept note for this piece of specific work form the PMA data cloud server archive via. https://www.pmadata.org/data/request-access-datasets.

